# Estimating the value of combination vaccines: a methodological framework

**DOI:** 10.1101/2025.09.05.25335206

**Authors:** Mark Jit, Allison Portnoy, Clint Pecenka, William P. Hausdorff, Christopher Gill

## Abstract

Combination vaccines combine several components in a single dose administration. They offer programmatic and public health advantages, particularly as vaccine schedules become increasingly crowded. They are often more expensive to develop and produce, which discourages manufacturer investment without clear market signals. Hence their benefits need to be captured with existing health economic evaluation reference cases used by decision-makers to guide vaccine investments. We propose that the value of combination vaccines can be captured through at least four domains: (i) reductions in tangible and intangible costs to caregivers; (ii) operational efficiencies to the health system; (iii) opportunity costs of vaccine schedule slots; and (iv) more streamlined vaccine schedules. We demonstrate the practicality of our framework by comparing the value of introducing a hypothetical vaccine to a crowded schedule as a standalone formulation, a replacement for a vaccine already in the schedule, or a combination product. The framework could also be applied to estimate the value of reducing the number of separate administrations needed for a standalone vaccine. Applying it in real-world situations could be facilitated by further data collection, particularly on collating results on the value of existing vaccines in the schedule, and estimating willingness-to-pay for fewer vaccine administrations.

**Key points for decision-makers:**

- Combination vaccines often have higher prices than stand-alone vaccines, so their value needs to be clearly established.
- Their value can be quantified in at least four domains (caregiver cost reductions, operational efficiencies, opportunity cost savings and streamlined schedules).
- The framework and hypothetical example presented here can be used to support appropriately valuing a combination vaccine.

## 1. Introduction

Vaccines are one of the greatest public health success stories of the past century, averting millions of deaths annually[1]. The pipeline of vaccines continues to grow, offering protection against both established and emerging pathogens[2]. However, as more vaccines are added to crowded vaccine schedules – particularly for infants and young children – healthcare providers and caregivers have raised concerns about the number of injections children receive and the overall complexity of the schedule[3].

Combination vaccines, which combine several components in a single dose administration, help to address these concerns. Widely-used examples include measles-mumps-rubella-containing vaccines (MMR or MMRV including varicella) and hexavalent vaccines (DTP-HepB-Hib-IPV, i.e., including antigens for diphtheria, tetanus, pertussis, Hepatitis B, *Haemophilus influenzae* type B, and polio), with others under development. While these combination vaccines could have programmatic and public health benefits, they face complex development, regulatory, manufacturing, and intellectual property challenges before they can be deployed[4,5]. These factors contribute to their generally higher procurement costs compared to the standalone vaccines they are replacing. For instance, incorporating inactivated poliovirus vaccine (IPV) to Serum Institute of India’s hexavalent vaccine increased procurement costs by 3-6 USD per series—up to 40% more than procuring standalone DTP-HepB-Hib and IPV vaccines [6].

Manufacturers are only incentivized to invest in developing combination products if they receive clear market signals that purchasers are willing to pay higher prices for these combinations. If a manufacturer faced losing a market because a country was considering replacing a standalone vaccine with a combination vaccine produced by a competitor, that could also incentivize it to develop a new combination even if a higher price wasn’t guaranteed. For dual market vaccines, the return on investment for manufacturers is driven by the high-income country market, where procurement decisions are informed by recommendations from bodies such as National Immunisation Technical Advisory Groups and Health Technology Agencies. These recommendations increasingly use value assessment frameworks that incorporate (among other factors) economic efficiency metrics such as cost-effectiveness and return on investment[7,8]. However, existing economic frameworks do not ascribe extra value of combination vaccines. Some vaccines (e.g. typhoid) are primarily purchased in low- and middle-income countries, which rely on recommendations from the World Health Organization, as well as funding from bodies such as Gavi, the Vaccine Alliance, and the Gates Foundation. However, none of these institutions express any preference for combination vaccines.

Hence, on the surface, combination vaccines with higher prices may appear less cost-effective than standalone vaccines. However, combination vaccines have several benefits – including improving programmatic efficiency, reducing healthcare visits and injection burden, and accommodating additional components in the vaccine schedule – that can be translated into standard economic outcomes such as gains in QALYs (quality-adjusted life years) or DALYs (disability-adjusted life years). Yet to date, no standard methodology exists to capture the benefits of combination vaccines within existing value frameworks.

To address this methodological gap, we propose a framework for systematically quantifying and incorporating the full value of combination vaccines into standard health economic evaluations. We demonstrate proof of principle for our approach using a stylised case study of a hypothetical new combination vaccine, and propose a research agenda to enable its routine application in future vaccine assessments.

## 2. Value framework

Standard health economic evaluations such as cost-effectiveness and benefit-cost analyses compare the value of an intervention (such as a new vaccine) to its comparator (such as no vaccine) based on the following inputs: (i) the incremental cost of the intervention (including both its procurement and its delivery, but subtracting health care cost savings and sometimes wider economic benefits such as productivity gains); and (ii) the health benefits of the intervention (which can be monetised based on individual or societal willingness to pay for health). We propose that the value of combination vaccines can be measured using these inputs through four domains:

### 2.1. Value of fewer vaccine administrations to the household

Caregivers incur transportation costs and lose time spent on productive labour to reach vaccination points. Beyond these tangible costs, there are also non-financial burdens: inconvenience, discomfort, and anxiety around multiple injections. Parents report wanting to minimise the number of clinic visits they make, and the number of injections their child receives at each visit[9]. Only a small minority of parents are comfortable with more than four injections per visit [3]. Empirical studies using discrete choice experiments (DCEs) and contingent valuation methods have quantified caregivers’ willingness to pay (WTP) to avoid additional injections in the United States[9–11], Japan[12], and China[13,14] (see Table X).

**Table 1.**
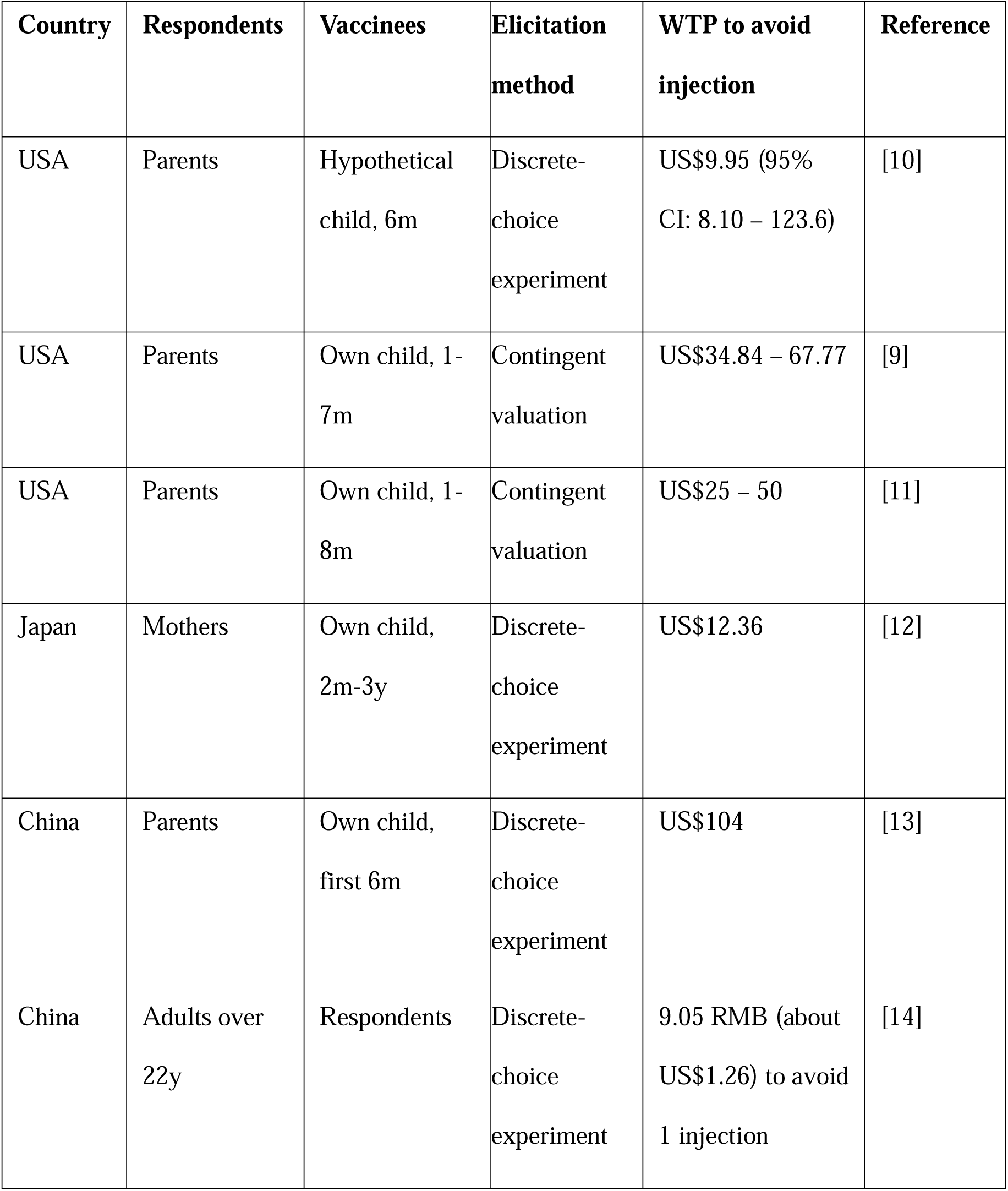
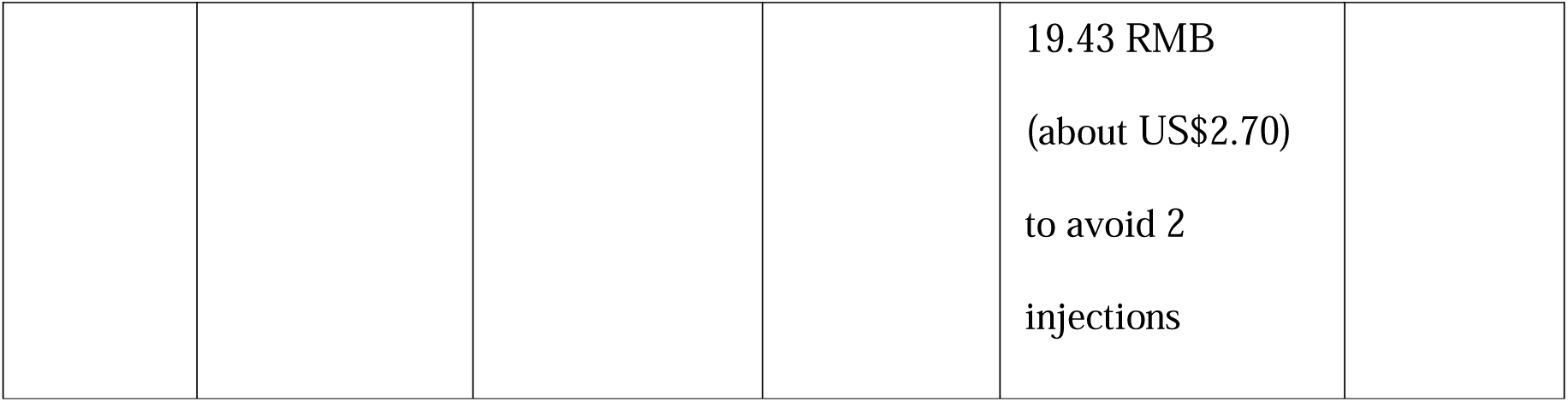
Willingness to pay (WTP) to avoid a vaccine injection, as measured in discrete choice experiments.

These costs were highest in one of the Chinese studies[13], although they were in the context of a high-income city (Shanghai) where parents often pay high prices for privately purchased imported vaccines. Some studies report that the level of discomfort changes with the number of administrations given (e.g., going from 0 to 1 administrations vs. 3 to 4 administrations); however, the direction of such change is not clear from existing studies[11,12,14]. There are no studies in low/middle-income settings.

### 2.2. Operational efficiencies to the health system

Each dose of vaccine administered carries costs to the health system. Firstly, by reducing the consumables needed, combination vaccines reduce the costs related to sustaining the vaccine supply chain at a country level. For example, measles-rubella and yellow fever vaccines are both lyophilized and require saline diluent for reconstitution. Each dose delivered therefore requires a vaccine vial, a diluent vial and two syringes (one to draw up and mix, one to administer). A combined measles-rubella-yellow fever vaccine reduces the number of administrations needed from three to two, saving two vials and two syringes. Besides the actual purchase costs of the consumables, more materials require greater costs for storage, cold chain, transportation, waste disposal and monitoring. Secondly, there are staff costs related to vaccine delivery at the clinic. A vaccination visit involves staff time to welcome the patient and caregiver, record their details, and introduce them to a vaccinator. Each vaccine administered separately during that visit also involves staff time to get the vaccine, administer it to the patient, counsel the patient and caregiver, and record the vaccination. Each administration also carries a risk of (suspected or actual) adverse events which need tracking, recording and patient management.

We used data from a study of the cost of pneumococcal, rotavirus and second dose measles vaccines in Zambia[15] to calculate that reducing the schedule by one administration could save 1.14 USD per infant in the birth cohort, or 0.73 million USD a year. Using a time and motion study in the United States [16], we found that reducing a dose could save 3.10 USD per infant in nurse time alone, or 11.2 million USD a year in total. Full calculations are shown in the Supplementary Appendix.

### 2.3. Opportunity cost of using a vaccine slot

Vaccination “slots” (timepoints in the schedule) are limited. Adding a new vaccine to an already crowded schedule may require displacing an existing vaccine or adding an additional visit—both of which involve trade-offs. In economic terms, placing a vaccine in a particular slot carries an opportunity cost. To use a real estate metaphor, the vaccine timeslots are “properties” and the vaccines given in those slots as “developments”. As in actual real estate, the location of these “properties” matters. For instance, if you are building a luxury hotel, you would typically choose a location with densely populated tourist attractions. However, there are many competing uses for real estate in that location, so building a hotel there will prevent the land from being used for much needed residential or commercial developments.

This highlights a dilemma in vaccine delivery. Vaccines can be delivered in a timeslot when no other vaccines are given (e.g., malaria vaccines at 5, 6, and 7 months), which requires the creation of a separate, costly visit. Alternatively, they can be delivered in a more established timeslot (such as 6, 10, and 14 months) where they compete with other vaccines for limited places. The first option has low opportunity costs but high administrative costs, while the latter involves the opposite trade-off. A combination vaccine would avoid both problems as it avoids needing to create an additional slot in the schedule.

The opportunity cost associated with using a particular slot is the value of the best (most cost-effective) vaccine to use in that slot. This can be measured in terms of avoided mortality and morbidity from disease prevented by vaccination, treatment cost savings, and any wider economic benefits that are admitted in the economic reference case of a jurisdiction (e.g., avoided productivity losses). Avoided mortality and morbidity can be monetized through standard economic evaluation methodology (e.g., value of avoided mortality in a cost-benefit framework, net monetary benefit in a cost-effectiveness framework).

### 2.4. More streamlined schedule

By reducing the number of visits needed, combination vaccines may improve vaccine coverage[17,18] and timeliness[19]. This would give greater overall protection from disease to the population. These benefits can hence be translated into health gains, and valued in a similar way to the above.

## 3. Hypothetical case study

We consider the scenario of a hypothetical single-dose vaccine (against disease X) that could be introduced into the schedule at either 4, 8, 12, or 16 weeks. We compare the economic value of three scenarios for vaccine introduction: (i) as a new standalone administration; (ii) as a replacement for an existing single-dose vaccine; or (iii) in a single formulation combined with the existing vaccine (see Table 2).

**Table 2.**
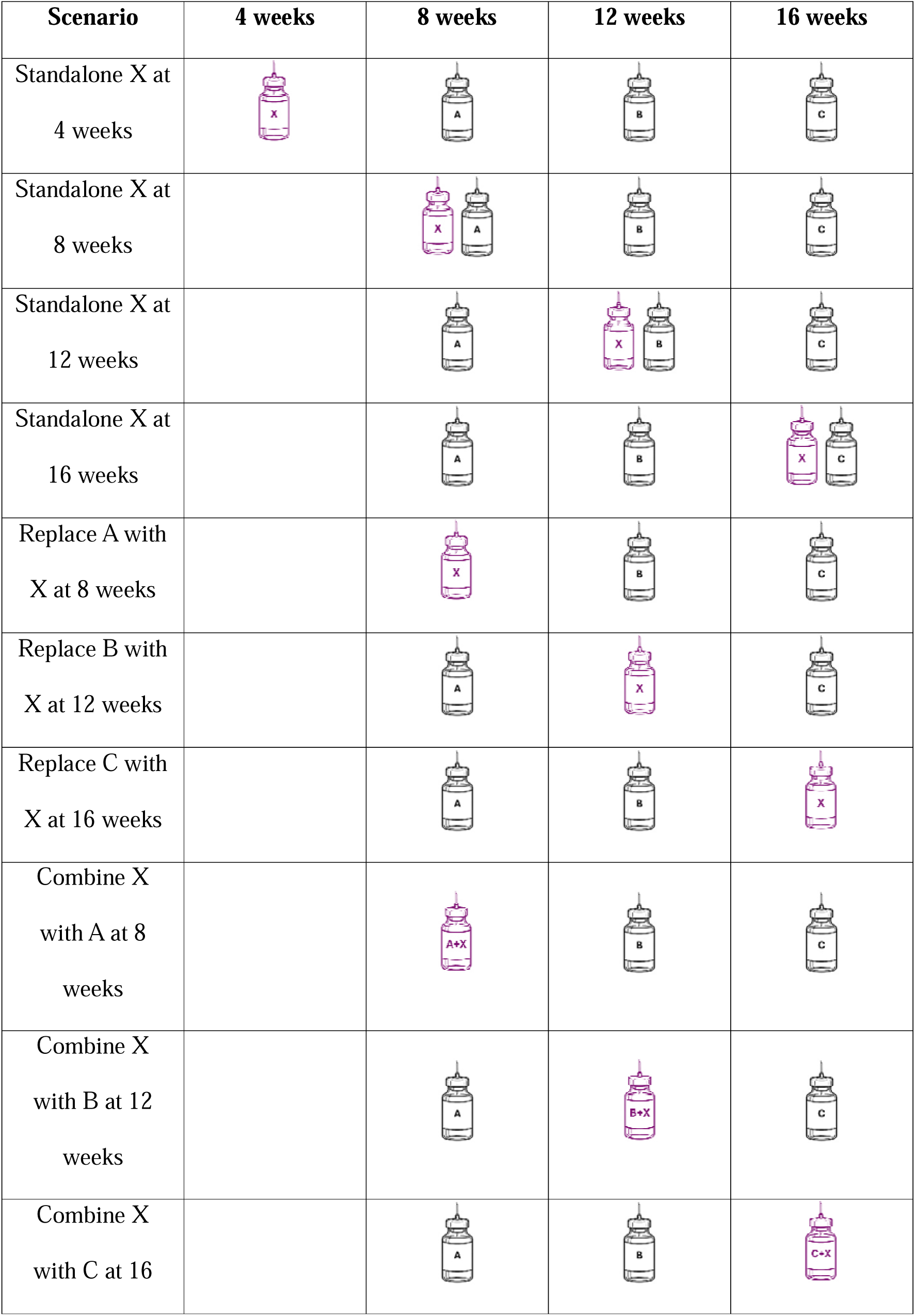
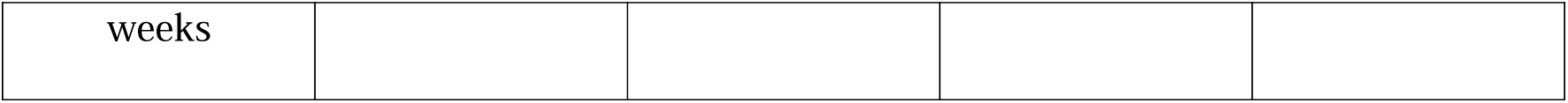
Scenarios showing all possibilities about vaccine X introduction to the schedule.

We also make the following assumptions: (i) the current schedule at those slots contains only three single-dose vaccines: vaccine A, vaccine B and vaccine C given at 8, 12 and 16 weeks respectively; (ii) the combination vaccine gives the same protection (i.e., has the same efficacy) as each of its individual components given as standalone vaccines.

The economic value of each scenario is determined in the following way, with the actual values assigned shown in Table 3:

**Table 3.**
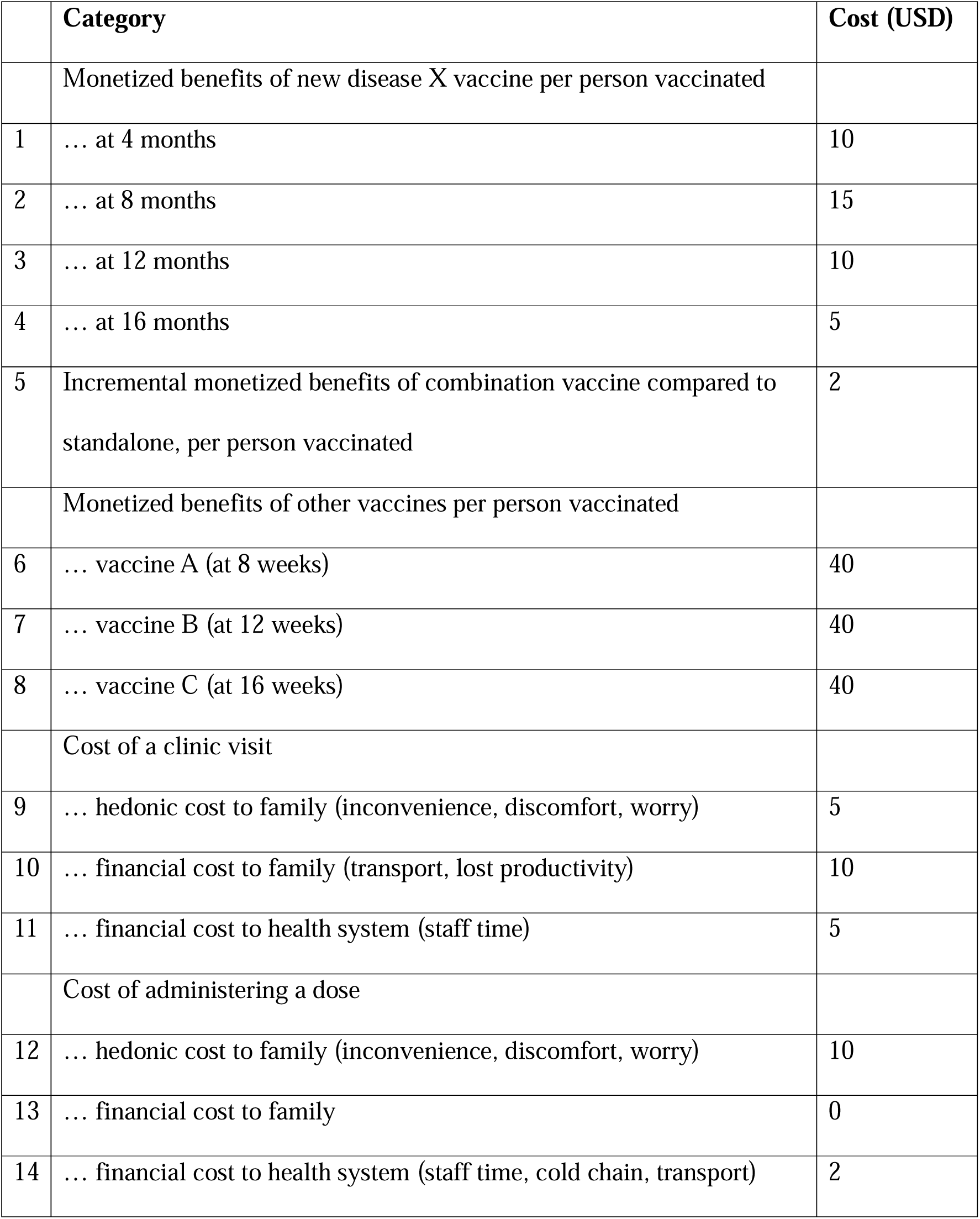
Hypothetical assumptions about economic costs and benefits around giving infant vaccines.

### Health benefits of disease X vaccine

The health gains (and hence value of disease avoided) of the new disease X vaccine depends on the timing of vaccination. We assume that disease incidence starts to rise at 4 weeks and peaks soon after. If the dose is given early (e.g., 4 weeks), then immunogenicity will be lower and protection will wane before the infant ages beyond the age of peak disease incidence. If it is given late (e.g., 16 weeks), then the infant will be unprotected over part of the duration of peak incidence. We also assume that a combination vaccine will be slightly more timely, and hence have slightly greater benefit.

### Health benefits of other vaccines

If introducing the new disease X vaccine displaces another vaccine administration from the schedule, this substitution leads to a loss of the health benefits of the displaced vaccine. The resulting health loss can be monetized, and for analytical purposes, this value can be allocated across the different administrations of a vaccine.

### Economic cost of vaccination-related activities

We assume that making a visit to a vaccination facility and giving a vaccine dose both impose costs to the health system and the vaccinee’s family. This includes hedonic costs to the family, i.e. costs of intangible attributes of the vaccination process such as causing inconvenience, worry, and discomfort. We also assume that giving more than four doses to a child per visit is not acceptable to the family.

### Calculating the value of a combination vaccine

By combining each element above, we can consider the net value of each option for introducing the new disease X vaccine, and at different ages. Based on these calculations (Table 4), if a combination vaccine did not exist, then the best value strategy would be to give it as an additional injection at age 8 months, and it would have a net value of 3 USD. Indeed, in this example it is the only strategy of using the vaccine as a standalone with a positive net value. If it could be given as a combination vaccine, then the best strategy would be to combine it with vaccine A at 8 months, and it would have a net value of 17 USD. The difference in value (14 USD) between the two best options is the value of a combination vaccine per person vaccinated, i.e., the additional cost that society could pay for a combination vaccine compared to a standalone.

**Table 4.**
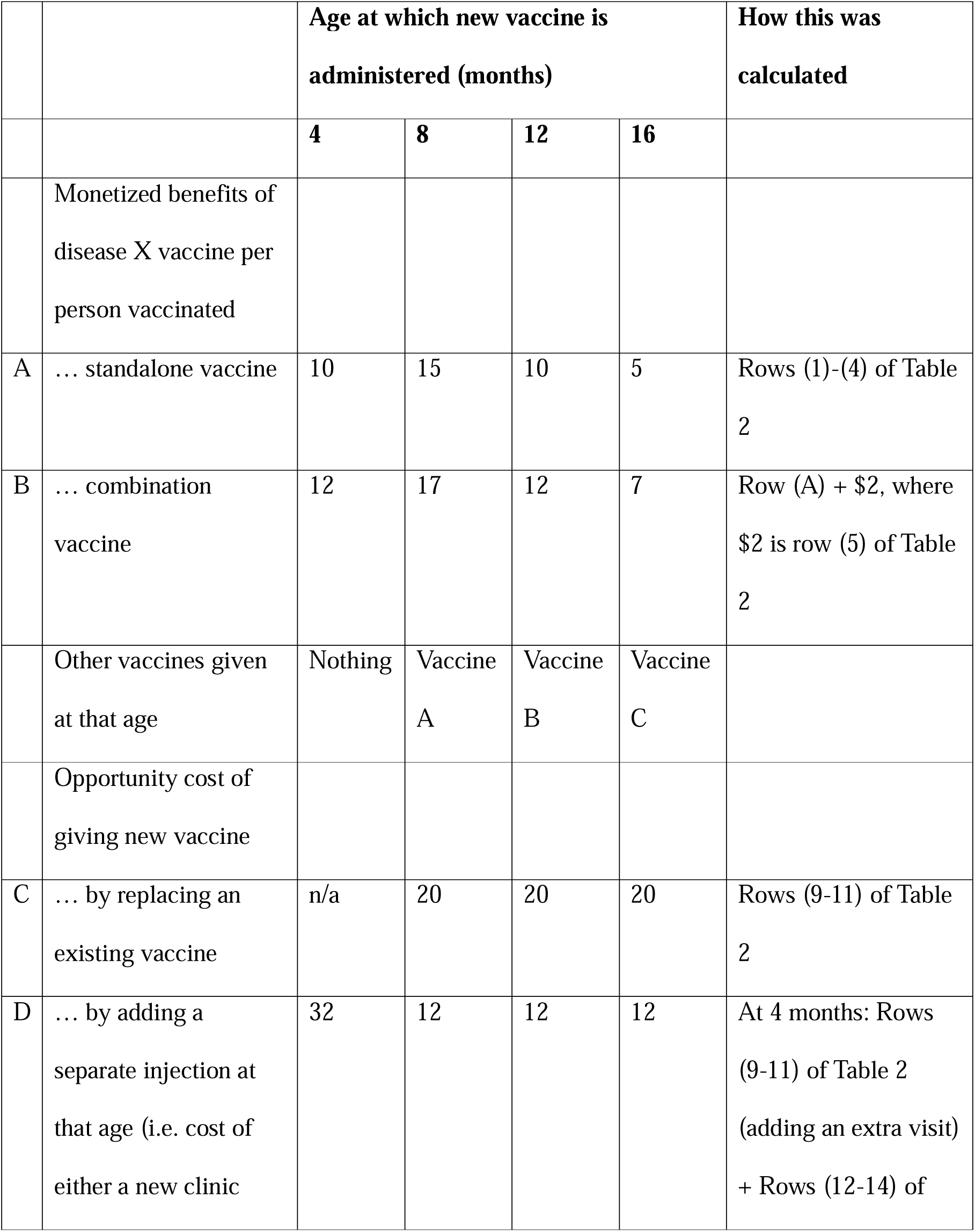
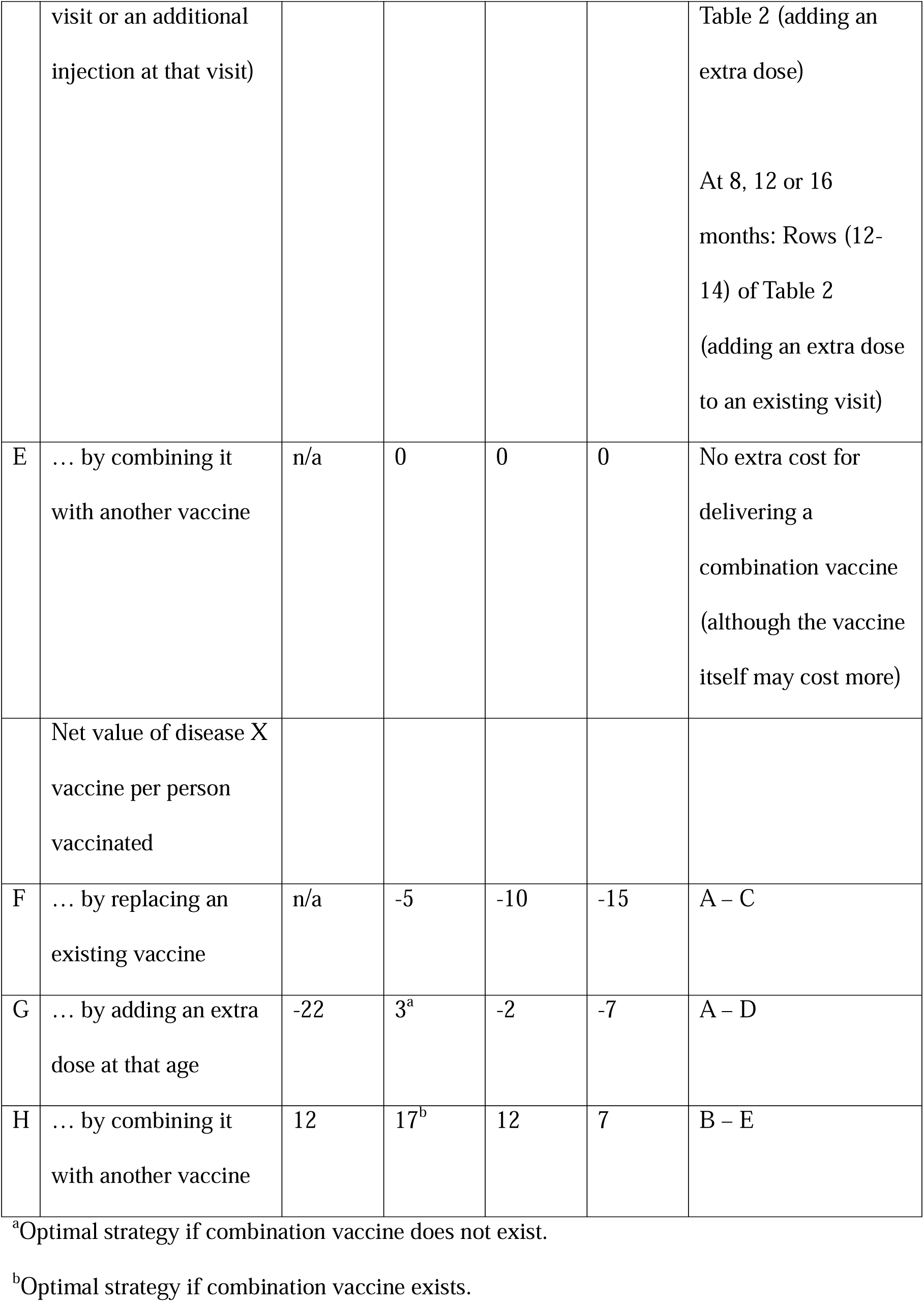
Value of a disease X vaccine under three scenarios (replacing an existing vaccine, giving it as an extra dose, giving it as a combination vaccine) and four possible target ages.

## 4. Discussion

Combination vaccines involve higher development and manufacturing costs due to their increased complexity. They will only be developed if their benefits to vaccinated populations justify these additional costs, allowing them to command higher prices than standalone vaccines. Our analysis has demonstrated how established methods in health economic evaluation can support a framework for valuing combination vaccines, by including assessment of the value of operational efficiencies to the health system, reductions in tangible and intangible costs to caregivers, opportunity costs of vaccine schedule slots, and more streamlined vaccine schedules. We have also shown that this conceptual framework is in principle straightforward to apply.

Our framework can be applied to value combination vaccines in three situations:

1. Reduced administrations: When combinations consolidate multiple components into a single injection, the framework quantifies the value of reducing the number of administrations.
2. Constant administrations with added components: When the number of administrations remains the same but new components are introduced, our framework captures the value of delivering greater protection (and health gains) without increasing the schedule burden.
3. Additional injections with new components: When vaccines combine several new components that require additional administrations, the framework evaluates whether the health gains justify the added delivery costs.

Additionally, while our framework was designed for combination vaccines, it also has utility in evaluating other strategies such as reducing the number of dosess of a single-component vaccine. For example, countries are considering simplifying their schedules for both human papillomavirus vaccine (from two doses to one)[20] and PCV (from three doses to two) [21], both to save procurement costs and to free up vaccine slots.

Combination vaccines may also have drawbacks which our framework can capture, even though we did not incorporate them in our hypothetical example. The first is reduced desirability. While most caregivers prefer to reduce the number of clinic visits and vaccine administrations their children receive, some may also have safety concerns around combining multiple components in a single administration [22]. This could be reflected in terms of reduced vaccine uptake. It could also be quantified through revealed preference studies as a hedonic cost if it results in increased caregiver anxiety. Another disadvantage of combination vaccines is reduced schedule flexibility. Countries uninterested in particular components in a combination vaccine (because of low burden for example) may be forced to introduced them anyway, if combination vaccines displace the market for single-component alternatives.

Hence economic evaluations of combination vaccines should ideally be multi-country studies that include settings which do not place high value on some of constituent components. A third disadvantage is that combination vaccines may compromise ideal timing in a schedule, as they require trade-offs between the optimal timing of each component. For instance, a malaria-hexavalent combination would require giving malaria vaccine at an earlier age to accommodate the hexavalent schedule, which may compromise its efficacy. However, earlier malaria vaccine administration may achieve higher coverage, and would avoid needing new vaccination visits at 5-7 months of age.

Our framework has two limitations. First, beyond the benefits of combination vaccines quantified in our analysis, combination vaccines are likely to offer other cost savings that are harder to quantify due to a lack of data. These include simplified supply chains, harmonized public messaging, streamlined training and record keeping[23], reduced needle stick injuries[24], and even shorter infant crying times. A simplified vaccine schedule could also contribute to reductions in greenhouse gas emissions associated with production, transport, and storage of vaccine doses. Also, by improving vaccine coverage, combination vaccines could improve health equity, since population groups with high disease burden are most likely to miss vaccine doses (and hence have the most to gain from coverage improvements).

Second, while applying our framework is straightforward, parameterising a real-world analysis requires estimates of key quantities needed in the analysis. One such quantity is the opportunity cost of vaccine slots. In many areas of health economic evaluation, estimating the opportunity cost of an intervention from first principles is challenging, as it requires evaluating a huge number of alternatives to identify and value the “next best alternative foregone”. For example, the opportunity cost of occupying a hospital bed would involve evaluating all the alternative patients who could have occupied that same bed[25]. In our hypothetical example, the opportunity cost of using a slot is measured by the health loss from removing an existing vaccine to make space for a new one. In practice however, removing an existing vaccine from the schedule is likely to be politically difficult. A more realistic scenario is that a future vaccine, despite its potential benefits, will never be introduced because all slots are already full. This still represents an opportunity cost, now framed in terms of vaccines that could be added in the future. Estimating this cost will require reliable estimates of the value of all possible vaccines that could occupy that slot. While such data exists in published cost-effectiveness studies, they still need to be systematically compiled to inform combination vaccine analyses.

Hedonic costs are another area needing more data collection. The financial and economic costs associated with clinic visits and doses are well documented[26]. However, the hedonic costs – such as inconvenience, worry, and discomfort experienced by children and their caregivers – are less well understood. DCEs have been used to measure such costs in a few countries, but more such data are needed, particularly in low- and middle-income countries. Estimates where doses are traded off against health rather than consumption would also be useful for analyses within an extra-welfarist framework (such as cost-effectiveness analysis).

## 5. Conclusion

Combination vaccines provide multiple programmatic and public health advantages that should be reflected in health economic evaluations to guide appropriate investment in their development and deployment. Targeted studies are needed to collect key parameters to inform such analyses.

## Statements and declarations

## Data availability

All datasets generated by this work are fully presented in the manuscript.

## Author contributions

MJ, AP and CG contributed to the study conception and design. Analysis was performed by MJ with input from all authors. The first draft of the manuscript was written by MJ and all authors commented on previous versions of the manuscript. All authors read and approved the final manuscript.

