## Supplementary Appendix for "Estimating the value of combination vaccines: a methodological framework"

***Estimation of health system costs saved by reducing the vaccine schedule by one dose in Zambia and the United States.***

***Zambia***^1^

By examining the immunisation costs categories in a micro-costing study of measles second dose (MCV2), pneumococcal conjugate (PCV) and rotavirus (RV) vaccination in Zambia, we find that the following categories would be impacted by reducing the number of doses:

- One-time investments: cold storage investments, tally sheets and under-5 cards, monitoring tools = 214,194 + 167,333 + 108,909 = 490,436 out of 1,161,337 (42% of total)
- Recurring costs: human resources, vaccine transport, cold storage, injection supplies = 2,634,548 + 941,715 + 199,958 + 135,872 = 3,912,093 out of 13,533,670 9 (29% of total)

Total number of doses given = 1 (MCV2) + 3 (PCV) + 2 (RV) = 6

Hence costs of each additional dose = 1/6 × (490,436 + 3,912,093) = 733,754

Total annualised costs for MCV2, PCV and RV = 14,695,007

This is 733,754/14,695,007 = 5% of total costs

Cost per infant in birth cohort = 22.89

Hence saving per infant in birth cohort by reducing one dose = 5% × 22.89 = 1.14

***United States***^2^

A time and motion study^2^ found that the total nurse time taken to administer a dose of vaccine = 1.7 (injection room) + 2.4 (examination room) = 4.1 mins

Hourly salary of a registered nurse (Jan 2025) = 45.42 (<https://nursa.com/salary/rn>)

Cost of nurse time per dose of vaccine = 4.1/60 × 45.42 = 3.10

Number of births in 2024 in USA = 3,622,673 (<https://www.cdc.gov/nchs/pressroom/nchs_press_releases/2025/20250423.htm>)

Total cost saved = 11,230,286
